# IgG1 pan-neurofascin antibodies identify a severe yet treatable neuropathy with a high mortality

**DOI:** 10.1101/2021.01.29.21250485

**Authors:** Janev Fehmi, Alexander J. Davies, R. Jon Walters, Tim Lavin, Ryan Keh, Alexander M. Rossor, Tudor Munteanu, Norman Delanty, Rhys Roberts, Dirk B□umer, Graham Lennox, Simon Rinaldi

**Author notes:** **Corresponding author: Dr Simon Rinaldi**, Nuffield Department of Clinical Neurosciences, University of Oxford, Level 6, West Wing, John Radcliffe Hospital, Oxford, UK. OX3 9DU.

## Abstract

Guillain-Barré syndrome and chronic inflammatory demyelinating polyneuropathy are umbrella terms for a number of pathologically distinct diseases involving disabling, immune-mediated injury to peripheral nerves. Current treatments involve non-specific immunomodulation. Prospective identification of patients with specific disease mechanisms should increasingly inform the use of more targeted disease modifying therapies and may lead to improved outcomes. In this cohort study, we tested serum for antibodies directed against nodal/paranodal protein antigens. The clinical characteristics of antibody positive and seronegative patients were then compared. Eight patients with IgG1-subclass antibodies directed against both isoforms of the nodal/paranodal cell-adhesion molecule neurofascin were identified. All developed rapidly progressive tetraplegia. Cranial nerve deficits (100% versus 26%), autonomic dysfunction (75% versus 13%) and respiratory involvement (88% versus 14%) were more common than in seronegative patients. The response to intravenous immunoglobulin, steroids and/or plasmapheresis was poor. Four patients received the B-cell-depleting therapy rituximab. After several weeks, these patients began to show progressive functional improvements, became seronegative, and were ultimately discharged home. Four patients who did not receive rituximab did not improve and died following the development of infectious complications and/or withdrawal of intensive care support. IgG1 pan-neurofascin antibodies define a very severe autoimmune neuropathy. We urgently recommend trials of targeted immunotherapy for this serologically-classified patient group.

## Introduction

Guillain-Barré syndrome (GBS) is characterised by flaccid limb weakness, supressed deep tendon reflexes, and a monophasic disease course reaching nadir within 4 weeks. Cranial nerve and autonomic dysfunction are common, and around 25% of affected individuals develop neuromuscular respiratory failure.[1] Demyelinating and axonal subtypes are defined by neurophysiology.[2] In chronic inflammatory demyelinating polyneuropathy (CIDP), disease activity and clinical progression continue for more than 8 weeks from onset.[3]

The node of Ranvier facilitates fast and efficient saltatory conduction along myelinated axons, which is reliant on the strict localisation of voltage-gated sodium channels and voltage-gated potassium channels at the node and juxtaparanode, respectively. This is ensured in part by cell adhesion molecules at the node (neurofascin-186 and gliomedin) and paranode (contactin-1, contactin-associated protein, and neurofascin-155).[4]

Pathology affecting the node, termed ‘nodo/paranodopathy’, has been linked to some forms of GBS,[5] in which anti-ganglioside antibodies capable of inducing complement-mediated nodal injury are found.[6] Recently, antibodies directed against nodal/paranodal proteins have been identified in atypical forms of CIDP.[4,7–12]

Herein, we describe eight patients with a very severe neuropathy associated with ‘pan-neurofascin’ IgG1-subclass antibodies and an atypical treatment response.

## Methods

From July 2017 to May 2020, we tested serum samples from 649 patients with suspected inflammatory neuropathies for IgG antibodies directed against nodal (neurofascin-186) and paranodal (neurofascin-155, contactin-1 and contactin-associated protein, Caspr1) cell-adhesion molecules, using a live, cell-based assay.[12] A standardised request form was used to collect clinical data. This study was approved by the NHS National Research Ethics Service Committee (South Central – Oxford A, 14/SC/0280). Further methodological details are given in the online appendix. The data that support the findings of this study are available from the corresponding author, upon reasonable request.

## Results

Overall, 46/685 patients with suspected inflammatory neuropathies (6.7%) were positive for nodal/paranodal IgG-class antibodies. These antibodies were not detected in 90 patients with other neurological diseases (20 with multiple sclerosis, 70 with antibody-positive CNS disorders) or in 120 healthy individuals. Seropositive patients consisted of 17 (2.5%) with antibodies against neurofascin-155 alone, 1 (0.15%) with monospecific neurofascin-186 antibodies, 11 (1.6%) with contactin-1 antibodies alone, and 9 (1.3%) with Caspr1 and/or contactin-1/Caspr1 complex antibodies. Eight patients (1.2%) had IgG antibodies which cross-reacted with both the nodal/axonal neurofascin-186 isoform, and paranodal/glial neurofascin-155 isoform (subsequently termed ‘pan-neurofascin’) (Figure 1A). These antibodies were exclusively IgG1, and IgG3 and/or IgG4 subclass antibodies with pan-neurofascin reactivity were not detected (Supplementary Figure 1). In contrast, using the same assays, 13/17 patients with neurofascin-155 (NF155) monospecific antibodies were IgG4 NF155 positive, and IgG1 NF155 was exclusively detected in only three cases. IgG2 and IgG3-subclass NF155 monospecific antibodies could occasionally be detected at lower intensities (Supplementary Table 1, Supplementary Figure 1). All eight pan-neurofascin positive sera, and all 17 NF155 positive sera, were negative for contactin-1 and Caspr1 antibodies. As the clinical associations of IgG1-subclass pan-neurofascin antibodies have not been reported, we sought to assess the characteristics of this serologically-defined cohort and compare these to seronegative patients and those with neurofascin-155 monospecific antibodies.

**Table 1.** Clinical features of patients with pan-neurofascin antibodies and comparison to neurofascin-155 and seronegative cohorts Contingency data were analysed using Fisher’s exact test and the 95% confidence interval (CI) of the odds ratio (OR) calculated by the Baptista-Pike method. Nadir mRs and CSF protein were compared by a two-tailed Kruskal-Wallis test with Dunn’s correction for multiple comparisons. Clinical data was available for all 8 panNF antibody positive patients, 15/17 NF155 patients, and 194/696 seronegative patients. Not all patients underwent MRI, and for some information on certain clinical features was not available, hence the lower denominators for some variables. GBS – Guillain-Barré syndrome, MRI – magnetic resonance imaging, mRs – modified Rankin scale, CSF – cerebrospinal fluid.

|  | **Pan-neurofascin**<br>**(n=8)** | **Neurofascin-155**<br>**(n=17)** | **Seronegative**<br>**(n=606)** | **OR**<br>**versus**<br>**NF155** | **95% CI** | **OR versus**<br>**seronegative** | **95% CI** |
| --- | --- | --- | --- | --- | --- | --- | --- |
| **Clinical diagnosis of GBS**<br>**(n, %)** | 5/8 (63%) | 3/15 (20%) | 38/185 (21%) | 6.7 | 1.1 - 34.5 | 6.5 | 1.6 - 24.9 |
| **Acute/subacute progression**<br>**(n, %)** | 8/8 (100%) | 7/15 (47%) | 56/184 (30%) | ∞ | 2.0 - ∞ | ∞ | 4.8 - ∞ |
| **Ataxia (n, %)** | 3/8 (38%) | 7/15 (47%) | 62/158 (39%) | 0.7 | 0.1 - 3.4 | 0.9 | 0.2 - 3.7 |
| **Tremor (n, %)** | 0/8 (0%) | 5/15 (33%) | 39/154 (25%) | 0 | 0 - 1.3 | 0 | 0 - 1.4 |
| **Neuropathic pain (n, %)** | 4/8 (50%) | 1/15 (7%) | 49/134 (37%) | 14 | 1.3 - 180 | 1.7 | 0.5 - 6.2 |
| **Cranial nerve palsy (n, %)** | 8/8 (100%) | 5/15 (33%) | 41/156 (26%) | ∞ | 3.3 - ∞ | ∞ | 5.7 - ∞ |
| **Autonomic dysfunction (n, %)** | 6/8 (75%) | 0/15 (0%) | 9/71 (13%) | ∞ | 6.1 - ∞ | 20.7 | 3.9 - 105.4 |
| **Respiratory involvement (n, %)** | 7/8 (88%) | 0/15 (0%) | 25/185 (14%) | ∞ | 9.9 - ∞ | 44.8 | 7.3 - 506.3 |
| **Nephrotic syndrome (n, %)** | 3/8 (38%) | 0/15 (0%) | 5/147 (3%) | ∞ | 1.9 - ∞ | 17 | 3.5 - 73.7 |
| **MRI Plexus/root abnormalities**<br>**(n, %)** | 2/4 (50%) | 2/7 (29%) | 15/53 (28%) | 6.7 | 1.1 - 34.5 | 6.5 | 1.6 - 24.9 |
| **nadir mRs >4 (n, %)** | 8/8 (100%) | 3/15 (20%) | 38/185 (21%) |  |  |  |  |
|  |  |  |  | **significant**<br>**v. NF155** | **p value** | **significant**<br>**v.**<br>**seronegative** | **p value** |
| **nadir mRs (median, range)** | 5.5 (5-6) | 4 (2-5) | 3 (1-5) | ** | 0.006 | *** | <0.001 |
| **CSF protein (g/L)**<br>**(median, range)** | 0.48 (0.34-0.62) | 1.65 (0.61-7.05) | 0.87 (0.18-6.0) | *** | <0.001 | * | 0.04 |

**Figure 1.**
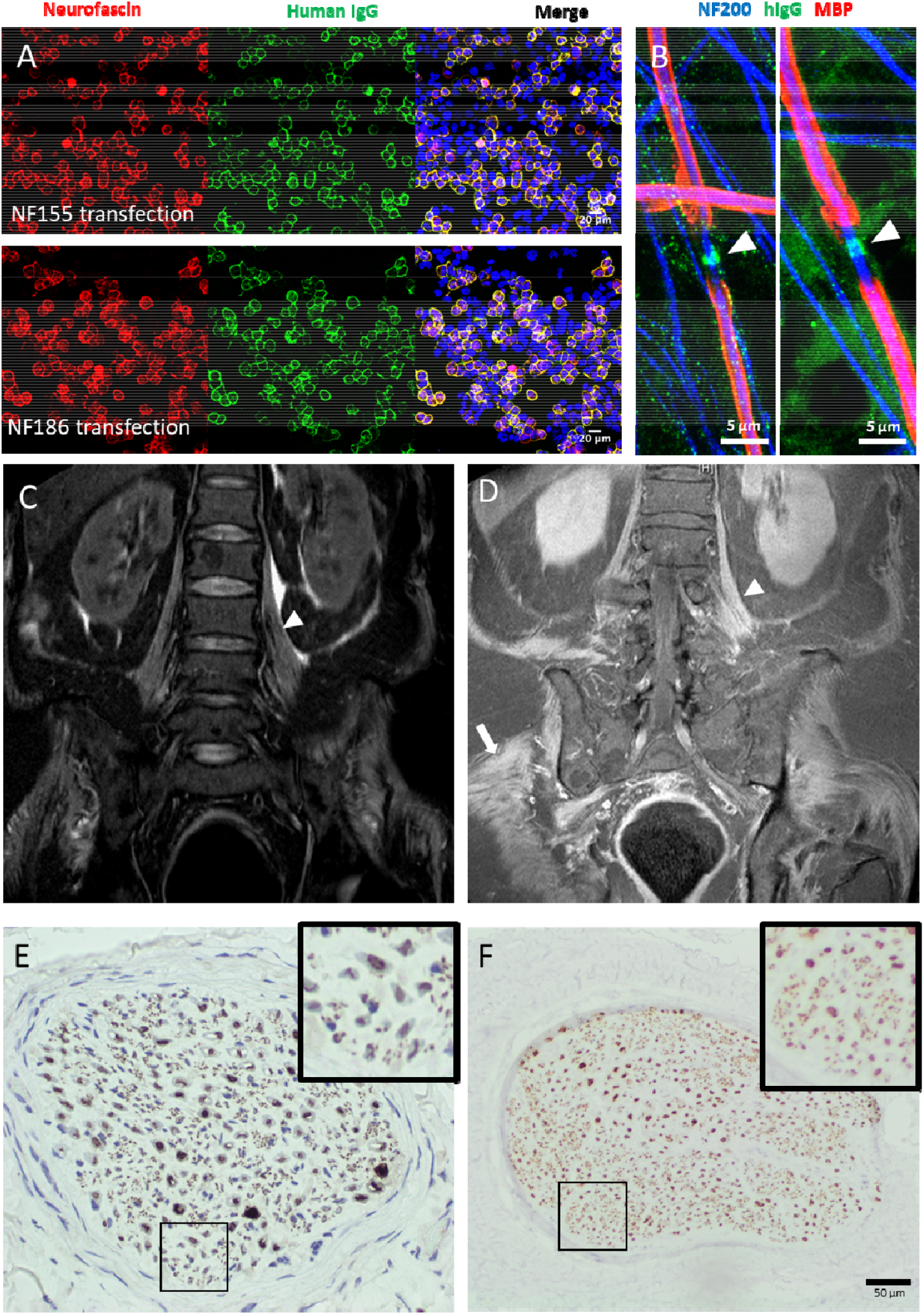
Serological, radiological and histological findings. **(A)** Cell-based assays using HEK293T cells transiently transfected to overexpress neurofascin-155 (upper panels) or neurofascin-186 (lower panels). Neurofascin (red) expression in the cell membrane is revealed by a commercial polyclonal antibody, and colocalises with human IgG (green) after exposure to acute-phase serum from the patients described in this series. **(B)** IgG (green) from two pan-neurofascin antibody positive patient sera (*left*, P1, *right*, P6) is deposited at the node of Ranvier (arrowhead) after exposure to myelinating co-cultures. Axons (Neurofilament-heavy, NF200, blue) were also observed weakly labelled with punctate IgG deposition in P1. Neither nodal or axonal labelling was observed in sera from healthy controls (data not shown). Myelin basic protein (MBP, red) defines the myelinated internode. **(C, D)** MRI of the lumbar spine, with coronal STIR **(C)** and post-contrast T1 **(D)** sequences, shows diffuse symmetric thickening and enhancement of lumbosacral plexus nerve roots (arrowhead), and enhancement of paraspinal, psoas, pelvic and proximal leg skeletal muscles (arrow). **(E)** Nerve biopsy from P6 stained for neurofilament (NFP) shows reduced numbers of NFP positive axons (more clearly seen in the inset panels), and dense patches of staining, consistent with axonal degeneration. “Near normal” NFP staining **(F)** is shown for comparison.

### Clinical features

Clinical details were available for all 8 pan-neurofascin antibody positive patients, 15/17 neurofascin-155 positive patients, and 194/606 seronegative cases. Pan-neurofascin antibody positive patients were all were very severely affected and had rapidly developed profound tetraplegia. Compared to seronegative patients, they were more likely to have presented following acute or subacute deterioration (odds ratio (OR) ∞, 95% confidence interval (CI) 4.8 - ∞), and to have received an initial clinical diagnosis of GBS (OR 6.5, 95% CI 1.6 - 24.9). Nadir modified Rankin scores (median 5.5, range 5-6) were significantly higher than those of both neurofascin-155 monospecific antibody positive (median 4, range 2-5, p=0.006) and seronegative patients (median 3, range 1-5, p<0.001, Kruskal-Wallis test with Dunn’s correction for multiple comparisons) (Table 1 and Supplementary Figure 2A). Indeed, the peak severity of the panNF patients was greater even than the subset of NF155 positive individuals with acute/subacute onset disease (median mRs 4, p<0.001, Mann-Whitney test). Cranial nerve palsies (100%), autonomic involvement (75%) and respiratory dysfunction (88%) were also more frequent (Table 1). Concurrently presenting nephrotic syndrome was more common than in seronegative patients (38% v 3%, OR 17, 95% CI 3.5 - 73.7), but less frequent than in patients with CNTN1 antibodies.[11] Ataxia (3/8), papilloedema (2/7), and neuropathic pain (4/8) were occasional features (Supplementary Table 2). Clinical vignettes for patients 1,5 and 6 are available on request. Patient 4 was described in a recent case report [13].

### Laboratory findings

During work-up of their neuropathy, two pan-neurofascin antibody positive patients were found to have an IgG-lambda paraprotein and were subsequently diagnosed with lymphoproliferative disorders (Hodgkin’s lymphoma and chronic lymphocytic leukaemia). A third was found to have a clonal urinary lambda light chain, without a serum paraprotein, that was not further investigated prior to his death. Three patients had features of nephrotic syndrome (peripheral oedema and hypoalbuminaemia) which had developed in parallel with their neuropathy, and, in two, urinary protein levels were analysed and nephrotic range proteinuria confirmed. All patients were otherwise negative for standard neuropathy screening bloods, including anti-GM1 and GQ1b-ganglioside antibodies. Cerebrospinal fluid (CSF) protein was either normal or only marginally elevated at presentation (median 0.51g/L, range 0.34-0.62), and significantly lower than that of both neurofascin-155 monospecific antibody positive (median 1.65 g/L, range 0.61-7.05, p<0.001) and seronegative patients (median 0.87 g/L, range 0.18-6, p=0.04, Kruskal-Wallis with Dunn’s correction for multiple comparisons) (Supplementary Figure 2D). CSF white cell counts were invariably normal (range 1-3/μL) with unremarkable cytology and flow cytometry. Using the cell-based assay (CBA), all patients had IgG1-subclass antibodies reactive against both neurofascin-155 and - 186 isoforms, and were negative on IgG2, IgG3 and IgG4 subclass-specific assays (Supplementary Table 1, Supplementary Figure 1A, C). End-point titres ranged from 1:400 to 1:6400 (Fig. 1A). Six of seven patients tested were also positive on a neurofascin ELISA, although the end-point titres were consistently lower than those obtained by CBA (Supplementary Figure 4). In 2 cases, positive results were only seen with one of the two neurofascin isoforms by ELISA, despite a clear pan-NF pattern on CBA. All eight pan-neurofascin positive and all 17 neurofascin-155 positive patients were negative for contactin-1 and Caspr1 antibodies. Five of the 7 pan-NF sera tested showed a characteristic and comparable nodal binding pattern in live, myelinating co-cultures (Fig. 1B), which was clearly different to that seen with neurofascin-155 monospecific sera (Supplementary Figure 1B, D). [14] No patients had IgG4 pan-neurofascin antibodies or developed these during follow up.

### Neurophysiology

Neurophysiological results were available for 6/8 patients. In one, the nerves were inexcitable when first assessed 2 weeks after onset. In 4/6, conduction slowing on initial studies was considered to indicate demyelination. However, 5/6 showed conduction block without temporal dispersion, suggestive of nodal pathology.[5] In three, follow up studies 3-4 weeks later revealed very reduced or unrecordable compound muscle action potentials and electromyographic findings consistent with severe axonal degeneration. Detailed neurophysiological results are given in Supplementary Table 3.

### Imaging

Four patients were examined by MRI. In one, symmetric enhancement and thickening of the lumbosacral plexus nerve roots, as well as enhancement of paraspinal, pelvic and proximal lower limb muscles, was observed. T2 hyperintensities of the brachial plexus and L5-S2 roots but no thickening or enhancement were seen in another (Fig. 1C,D).

### Histology

Nerve biopsy was performed in 2 patients. In both cases this demonstrated axonal loss, without any features of cellular infiltration, inflammation, segmental demyelination, amyloid or vasculitis (Fig. 1E, Supplementary Figure 3). Electron microscopy was performed in one patient and did not show any evidence of paranodal retraction/detachment. One patient had a necrotising myopathy on muscle biopsy with a normal creatine kinase.

### Treatment response and outcome

All patients received intravenous immunoglobulin (IVIg) 2g/kg over 5 days. In 6 cases this was not associated with any perceptible benefit. In 2 there was a minor and/or transient neurological improvement. Six patients received at least 1 cycle of plasma exchange (PLEx), with 3 patients showing slight but non-sustained neurological recovery. Further detail and physician reported assessments of response are given in Supplementary Table 4. Four patients died. One suffered a cardiorespiratory arrest 8 days after presentation, and in the absence of recovery of cortical function, ventilatory support was withdrawn 10 days later. In another, recurrent pulmonary infections and the absence of any neurological recovery after IVIg and 2 cycles of PLEx led to the withdrawal of ventilatory support on day 108. A third declined artificial ventilation, having failed to respond to steroids, IVIg, PLEx and cyclophosphamide, and died on day 93. Most recently, intensive care and mechanical ventilation was not deemed appropriate for one patient in the context of the COVID-19 pandemic. He was SARS-CoV-2 PCR negative, but developed increasing breathlessness and tachypnoea on day 12, and died 48 hours later. In both patients with nephrotic range proteinuria, this was still apparent when last measured shortly before their death.

The remaining 4 patients received rituximab (1g repeated after 2 weeks) several months into their illness, following no or minimal and transient responses to other therapies. In 2 patients, this was given with combination chemotherapy for newly diagnosed haematological disorders. Thereafter, all 4 rituximab-treated patients began to show progressive and sustained functional improvement, regained independent mobility, and were ultimately discharged home, often via a rehabilitation facility. All rituximab treated patients showed improvements of at least 2 points on the modified Rankin scale within 6 months of rituximab treatment, and one became asymptomatic by 12 months (Supplementary Figure 2B). In the one rituximab-treated patient with nephrotic syndrome, the serum albumin normalised in parallel with neurological improvement. In all four patients, neurofascin antibodies were negative when retested 4-11 months after rituximab. One of these 4 patients, having returned almost to his baseline (mRs=1, minimal residual symptoms) had a return of motor and sensory symptoms, approximately 18 months after receiving a single cycle of rituximab. He developed marked arm weakness, and became immobile over a few weeks. He redeveloped hypoalbuminaemia in parallel. At this stage pan-NF antibodies were again positive, though at low titre (1:100 and 1:200). Again, IgG1 was the dominant subclass. He was retreated with steroids and a second cycle of rituximab, and again improved, regaining independence.

## Discussion

This is the first reported series of patients with IgG1-subclass, pan-neurofascin antibodies. These antibodies are shown to be associated with an extremely severe and rapidly progressive neuropathy.

To date, antibodies directed against nodal and paranodal cell adhesion molecules have largely been described in patients diagnosed with ‘atypical’ chronic inflammatory demyelinating polyneuropathy (CIDP). In this context, IgG4 antibodies have been reported as pathologically important markers of a clinically distinct CIDP sub-group, whereas patients with only non-IgG4 subclass antibodies were found to be indistinguishable from seronegative individuals.[15] In our series, patients with IgG1-subclass pan-neurofascin antibodies, who had significantly different clinical features and disease severity compared to identically identified seronegative controls, were more likely to have received a clinical diagnosis of GBS.

In the 5 pan–neurofascin patients in our series diagnosed with GBS, there was no clinical basis to reclassify their neuropathy as CIDP. Three showed 1 or 2 transient fluctuations after treatment, but all within 8 weeks of onset, and 2 died inside 4 weeks.[16] The three other patients met the clinical and electrodiagnostic criteria for definite CIDP,[3] yet their neuropathy was probably not demyelinating.

At present, the distinction between GBS and CIDP can only be confidently drawn when ongoing deterioration is seen more than 8 weeks after onset.[16] This criterion is of little to no use in informing therapeutic decisions during the early phases of a rapidly progressive, but potentially chronically persistent, autoimmune/inflammatory neuropathy. As the immuno-pathological process in GBS is typically conceptualised as monophasic and short-lived, a diagnosis of GBS provides little impetus to give immunomodulatory therapy outside of the acute phase.

The improvements after rituximab seen here occurred several months into the illness, after poor responses to IVIg and/or plasma exchange. Our observations suggest that a more persistent autoimmune response can potentially drive ongoing axonal loss, and prevent recovery, in patients whose initial presentation nonetheless resembles GBS, and in whom clinical deterioration greater than 8 weeks from onset is not always apparent, preventing a diagnosis of CIDP. Serological results and biomarkers of ongoing peripheral nerve injury may in future prove to have greater utility in guiding these treatment decisions and may ultimately supersede clinical categorisation. The earlier use of potent immunotherapies with longer durations of action in such patients may be more effective in reducing long-term disability.

In this observational study, patients received multiple different therapies at different time-points. We cannot, therefore, provide any clear evidence on the efficacy of any particular therapy in this setting. It is possible that the 4 patients who survived long enough to be treated with rituximab would have started to improve even if this therapy had not been given. Future trials should address whether the earlier use of targeted immunotherapy in such serologically defined cohorts could ameliorate the very severe disease course seen here.

In 238 patients with clinical diagnoses of GBS, across 3 different cohorts, we have now detected pan-neurofascin antibodies in 5 (2.5%, 95% CI 0 to 5.3%). A more accurate estimate of the frequency of these and other autoantibodies, and their prognostic value, in patients with GBS-like presentations is now a planned component of the large-scale International GBS Outcome Study (IGOS).[17,1].

Cancer is rarely associated with GBS/CIDP,[18] and onconeural antibodies have not previously been identified. The frequency of IgG-lambda paraprotein associated lymphoproliferative disorders and solid organ malignancy in this cohort suggest that pan-neurofascin antibodies may be responsible for some such cases.

Antibodies against the paranodal isoform of neurofascin-155 have been linked “atypical CIDP”. Neurofascin-155-monopecific seropositive individuals are often younger males with predominantly distal weakness, sensory ataxia, and tremor. Neurofascin-155 autoantibodies are predominantly of the non-complement-fixing IgG4 subclass and the response to IVIg is typically poor.[8] Antibodies against nodal isoforms of neurofascin (140/186) have also been described in five patients with CIDP.[10] Again, 4/5 patients had predominantly IgG4-subclass antibodies, but 3/4 responded well to IVIg. The remaining patient had IgG3-subclass antibodies and did not respond to IVIg, but improved after steroids and plasma exchange. Similar to our cohort, 2/5 were found to have nephrotic syndrome. This complication appears less common than in patients with contactin-1 antibodies,[11] and may be explained by the expression of neurofascin-186 by glomerular podocytes as well as neurons.[19] A further five severely affected neurofascin IgG3 seropositive patients with poor response to standard therapies have recently been described. In all but one case, antibodies cross-reacted with both neurofascin 140/186 and neurofascin 155 in cell-based assays, as in our cohort. One patient treated with rituximab subsequently improved, as did another, apparently spontaneously, starting 3 months into his illness.[20–22] The patients with IgG3 neurofascin-140/186 and pan-neurofascin antibodies previously reported seem most similar to our cohort. The differences in the pan-neurofascin antibody-subclass distribution detected in our study may be due to technical factors. The secondary antibodies used here were shown to recognise recombinant human IgG of the relevant subclass, did not cross react with any of the other subclasses (Supplementary Figure 4), and were all detected with the same mouse tertiary antibody. However, we used cell-based assays to determine subclass, given their increased sensitivity for pan-NF antibodies, whereas most other studies used ELISA.

In summary, the observations herein provide further rationale for nodal/paranodal antibody testing in GBS-like presentations. They highlight the importance of testing against *both* glial and neuronal neurofascin isoforms (to distinguish pan-NF from NF155 monospecific antibodies) and determining the IgG subclass, as this may also influence the clinical phenotype and response to treatment. We believe that such testing should increasingly form part of the routine diagnostic process, and that the identification of a distinct, serologically-defined disease subtype should prompt consideration of more targeted immunotherapy. The possible benefit of rituximab reported here should be evaluated in a well-conducted clinical trial, the design of which must consider the potentially grave outcome in pan-neurofascin antibody-positive patients receiving only standard therapies.

## Supporting information

STROBE

JF Disclosure form

GL Disclosure form

DB Disclosure form

RR Disclosure form

TM Disclosure form

AMR Disclosure form

RK Disclosure form

RJW Disclosure form

AJD Disclosure form

TL Disclosure form

ND Disclosure form

SR Disclosure form

## Data Availability

The data that support the findings of this study are available from the corresponding author, upon reasonable request.

## Acknowledgements

The “near normal” nerve biopsy image was kindly supplied by Dr Monika Hofer, Consultant Neuropathologist, John Radcliffe Hospital, Oxford.

## Author contributions

JF wrote the initial draft of the manuscript and performed the immunoassays. AD contributed to the initial draft and revised and edited future iterations for intellectual content. He also performed the co-culture assays. RJW, TL, RK, AMR, TM, ND, RR, DB and GL all identified and assessed patients, collected clinical data, and revised the manuscript for intellectual content. SR conceived the study, helped to analyse the clinical data and revised the manuscript for intellectual content. All authors approved the final version for submission.

## Abbreviations

Caspr: contactin-associated protein (1)
CBA: cell-based assay
CIDP: chronic inflammatory demyelinating polyneuropathy
ELISA: enzyme-linked immunosorbent assay
GBS: Guillain-Barré syndrome
Ig: immunoglobulin
IVIg: intravenous immunoglobulin
NF: neurofascin
OR: odds ratio
CI: confidence interval
PLEx: plasma exchange

