## Supplementary material for "IgG1 pan-neurofascin antibodies identify a severe yet treatable neuropathy with a high mortality"

#### Supplementary methods

**Nodal/paranodal antibody cell-based assays**

All sera screened were initially screened using a live, transiently transfected cell-based assay (CBA), following previously described methods with slight modification.[1] Human embryonic kidney 293T ("HEK") cells were plated at a density of 75,000 cells/13mm coverslip, and mono-transfected with either neurofascin-155 (NF155, *NFASC*) (RC228652, Origene), neurofascin-186 (NF186, courtesy of Jerome Devaux, University of Marseille), or co-transfected with contactin-1 (*CNTN1*, EXA1153-MO29 Genecopoeia, Maryland, US) and contactin-associated protein ('1') (*CASPR1*, EXMO417-MO2 Genecopoeia, Maryland, US), diluted in Jet-PEI transfection reagent (101-10; Polyplus). Patient sera were screened using a dilution to 1:100 in DMEM/1% bovine serum albumin (BSA) for incubation with neurofascin transfected cells, then titred out to 1:6400. Co-incubation with commercial chicken anti-neurofascin primary antibody, (1:1000) (Cat no. AF3235; R&D Systems, Bio-Techne) was used to confirm successful transfection and to assess for co-localisation with any bound human IgG. Secondary antibody incubation was with goat anti-human IgG-Fc specific-Alexa Fluor 488 (1:750) (Cat no. H10120; Life Tech) and goat anti-Chicken Alexa Fluor 546 1:1000 (Cat no. A11040; Life Tech). To determine antibody subclass unconjugated mouse anti-human IgG subclass 1-4 antibodies were used at 1:100 (Cat nos. I2513, I25635, I7260 I7385; Sigma-Aldrich, Merck) with the fluorescently tagged tertiary antibody goat anti-mouse Alexa Fluor 488 (1:750) (Cat no. A11029; Life Tech).

**ELISA**

ELISA was performed as previously described.[2] In brief, Nunc Maxisorp ELISA plates (Fisher Scientific) were coated with either human recombinant neurofascin-155 (NF155) (8208-NF, R&D systems), NF186 (TP329070, OriGene Technologies) or CNTN1 (10383-H08H, Sino Biological Inc) at 1 μg/ml. Blank wells were incubated with carrier only (PBS). After overnight incubation at 4°C, the coating solution was tipped off and wells were blocked with 5% milk for 1h at room temperature. Sera were incubated for 1h at room temperature, and initially screened at 1:100 dilution in 5% milk. Following a wash step through 5 changes of PBS, anti-human IgG (Fc-specific) peroxidase-conjugated anti-human IgG (A0170, Sigma) was applied at 1:3000 in 5% milk and incubated for 1h at room temperature, then washed as before. The detection reaction was performed using o-Phenylenediamine dihydrochloride (OPD, Sigma), applied for 20 minutes in the dark, then terminated with 4M $H_2SO_4$. Optical densities were measured at 492nm. Wells with ODs greater than 0.1 above uncoated control wells were considered positive. The end-point titre of positive samples was then assessed by serial doubling dilution from 1:100 to 1:6400. The end-point titre was defined as the highest dilution with an OD greater than 0.1 above the uncoated control. To assess the specificity of the subclass specific secondary antibodies used in the CBA, recombinant human IgG subclass proteins (HCA049, HCA50, HCA178, HCA193, HCA194, HCA195, Bio-Rad) were coated as above, and then exposed to the unconjugated mouse anti-human IgG subclass 1-4 antibodies for 1h at room temperature. Following a wash step, a HRP anti-mouse-IgG antibody (A4416, Sigma) was applied at 1:3000 for 1h, washed off, and binding detected as before.

**Collection of clinical data**

A request form (over page) was sent to all clinicians submitting samples for nodal/paranodal antibody testing and initial information was requested before test results were known. Clinical data was obtained from all 8 patients with pan-neurofascin antibodies, 15/17 with neurofascin-155 antibodies, and 194/606 seronegative individuals. Two attempts were made to collect missing data by re-contacting referring clinicians directly. All patients without clincal data returns following these efforts were omitted from the subsequent analysis. Follow up times varied from 3 years to 2 months for the antibody positive cohort (average 19 months), with the shortest follow up for the surviving pan-neurofascin patient being 10 months. Nodal/paranodal and seronegative neuropathy patients seen in Oxford were recruited to an observational study. This study was approved by the NHS National Research Ethics Service Committee (South Central – Oxford A, 14/SC/0280). The use of de-identified clinical data from other patients to audit the relative value of available and novel methods to determine autoantibodies, based on the gold standard of immune-mediated clinical phenotypes, was approved by the Oxford University Hospitals NHS Trust clinical audit leads (ID 5106).

**Analysis of clinical associations**

Contingency data for the presence or absence of 10 core clinical features (diagnostic category, onset/progression, ataxia, tremor, neuropathic pain, cranial nerve palsy, autonomic dysfunction, respiratory involvement, nephrotic syndrome, and MRI abnormalities) were analysed by Fisher's exact test without family-wise correction for error rate. Point estimates of the odds ratio are reported, with 95% confidence intervals calculated by the Baptista-Pike method. The widths of the intervals have not been adjusted for multiplicity and as such the inferences drawn may not be reproducible. The nadir modified Rankin score and CSF protein level at diagnosis were compared by the Kruskal-Wallis test with Dunn's correction for multiple comparisons.

**Preparation and immunofluorescence of myelinating co-cultures**

Sera were assessed for topographical binding using myelinated co-cultures. These were generated using sensory neurons derived from induced pluripotent stem cells (iPSC) according to a previously described protocol.[3] iPSCs were differentiated into neurons over 10 days, and myelinated with rat Schwann cells after four weeks to establish mature myelinated co-cultures. For immunolabelling these were incubated with the patient's sera, diluted at 1:100, then fixed with 2% PFA before incubation with secondary antibody goat anti-human IgG AF488 (Cat no. A11013: ThermoFisher Scientific), and permeabilised with ice-cold methanol before incubation with secondary antibodies chicken anti-Neurofilament 200 (1:10,000) (Cat no. 4680; Abcam) and rat anti-myelin basic protein (1:500) (Cat no. 7349; Abcam). Tertiary antibody incubation was with biotinylated goat anti-chicken IgY (1:500) (Cat no. BA9010; Vector lab), goat anti-rat IgG Alexa Fluor 546 (1:1000) (Cat no. A11081 Life Tech) and Streptavidin pacific blue (1:500) (Cat no. S11222 Life Tech). Fluorescence images of IgG nodal labelling in myelinating cultures were acquired on a laser scanning confocal microscope (LSM 700, Zeiss) at x63 with 2x digital zoom using 405, 488 and 555 nm lasers. 10-15 z-sections at 0.5 μm interval were exported as maximum projection images. Brightness and contrast were adjusted for presentation.

### Request Form for Paranodal / Nodal Antibody Testing

**Clinical Data**

Date of neuropathy onset: Age at diagnosis:

☐ Prodromal illness/trigger (please specify):
Start date for prodrome/trigger:

Initial diagnosis
☐ GBS ☐ Typical CIDP ☐ MMN
☐ Atypical CIDP / Other (specify):

Current diagnosis
☐ GBS ☐ CIDP ☐ MMN
☐ Atypical CIDP / Other (specify):

**If the current diagnosis is GBS, CIDP, or MMN please answer the following (tick all that apply):**

Clinical course
☐ Relapsing-remitting ☐ Progressive ☐ Monophasic

Onset / progression
☐ Acute (<4 weeks) ☐ Subacute (4-8 weeks) ☐ Chronic (>8 weeks)

Weakness (Yes/No) (Tick all that apply)

☐ Arms
☐ Proximal ☐ Distal
☐ Asymmetric ☐ Symmetric

☐ Legs
☐ Proximal ☐ Distal
☐ Asymmetric ☐ Symmetric

Sensory deficit (Tick all that apply)

☐ Arms
☐ Vibration ☐ Pinprick ☐ JPS

☐ Legs
☐ Vibration ☐ Pinprick ☐ JPS

☐ Ataxia ☐ Tremor ☐ Neuropathic pain 0 /10 Severity (1-10)

Reflexes
☐ Absent ☐ Decreased ☐ Normal ☐ Brisk

☐ Cranial nerve involvement (specify)

☐ Autonomic involvement (please specify)

Respiratory involvement ☐ Current ☐ Previous ☐ None

Evidence of nephrotic syndrome
☐ Proteinuria (level: ) ☐ Oedema ☐ Not assessed
☐ Hypoalbuminaemia (nadir level: g/L) ☐ None

Severity
Modified Rankin score (at nadir): 1 ☐ 2 ☐ 3 ☐ 4 ☐ 5 ☐ 6 ☐

#### Investigations

☐ CSF (at diagnosis) Date:

Protein: g/L WCC: RCC:

OCBs: (select from drop down menu) Other:

Neurophysiology

Overall impression: ☐ Demyelinating ☐ Axonal ☐ Mixed Other (specify)

☐ Motor involvement (describe core features):

☐ Sensory involvement (describe core features):

Other Antibodies

Gangliosides ☐ Positive ☐ Negative ☐ Not done

Anti-MAG ☐ Positive ☐ Negative ☐ Not done

Paraprotein ☐ Positive ☐ Negative ☐ Not done

☐ IgG ☐ IgM ☐ IgA ☐ Kappa ☐ Lambda

Level: g/L

Imaging

MRI lumbar roots ☐ Abnormal ☐ Normal ☐ Not done

Specify:

#### Treatment and Outcome

| | Trialled | | Response: Good | Partial | None | Worse |
|---|---|---|---|---|---|---|
| **IVIg** | Yes ☐ | No ☐ | ☐ | ☐ | ☐ | ☐ |
| **Steroids** | Yes ☐ | No ☐ | ☐ | ☐ | ☐ | ☐ |
| **Plasma Ex.** | Yes ☐ | No ☐ | ☐ | ☐ | ☐ | ☐ |
| **Other** (specify) | | | ☐ | ☐ | ☐ | ☐ |
| **Other** (specify) | | | ☐ | ☐ | ☐ | ☐ |

#### Current Disease Activity

1.Cure: ≥5 years off treatment

- ☐ A. Normal examination
- ☐ B. Abnormal examination, stable/improving

2. Remission: <5 years off treatment

- ☐ A. Normal examination
- ☐ B. Abnormal examination, stable/improving

3. Stable active disease: ≥1 year, on treatment

- ☐ A. Normal examination
- ☐ B. Abnormal examination, stable/improving

4. Improvement: ≥3 months <1 year, on Treatment

- ☐ A. Normal examination
- ☐ B. Abnormal examination, stable/improving

5. Unstable active disease: abnormal examination with progressive or relapsing course*

- ☐ A. Treatment naïve or <3 months
- ☐ B. Off treatment
- ☐ C. On treatment

Modified Rankin score (at best post treatment):

0 ☐ 1 ☐ 2 ☐ 3 ☐ 4 ☐ 5 ☐ 6 ☐

| | Dominant subclass | | Subclasses detected | |
|---|---|---|---|---|
| | PanNF (n=8) | NF155 (n=17) | PanNF (n=8) | NF155 (n=17) |
| None | 0 | 1 | - | - |
| IgG1 | 8 | 5 | 8 | 12 |
| IgG2 | 0 | 0 | 0 | 8 |
| IgG3 | 0 | 0 | 0 | 1 |
| IgG4 | 0 | 11 | 0 | 13 |

*Supplementary Table 1 - Subclass distribution of pan-neurofascin versus neurofascin-155 monospecific antibodies*

In all cases, IgG1 was the only subclass detected for pan-neurofascin antibodies. In contrast, with neurofascin-155 monospecific antibodies, IgG4 was most often the dominant subclass detected, but IgG1 and to a lesser extent IgG2 were also frequently observed.

| Decade of age of onset / Gender | Clinical | | | | | | | | | Investigations | | | | Treatment (response) | | | | | Outcome |
|---|---|---|---|---|---|---|---|---|---|---|---|---|---|---|---|---|---|---|---|
| | **Clinical Diagnosis** | **Motor/ Sensory** | **CN palsy** | **Autonomic** | **Ataxia** | **Pain** | **Associated conditions** | **ITU / MV** | **Nadir mRS** | **Para-protein** | **NCS (Overall impression)** | **Nerve biopsy** | **Anti-NF155/ 186 Titre (CBA)** | **Steroids** | **IVIg** | **PLEx** | **RTX** | **Other** | |
| P1 ($7^{th}$ / F) | GBS | M&S | ✓[a] | ✓ | × | ✓ | Nephrotic syndrome | ✓ | 6 | ND | Inexcitable | × | 1:6400/ 1:3200 | × | ✓ (red) | ✓ (red) | × | × | Death (Day 110) |
| P2 ($7^{th}$ / F) | CIDP | Motor only | ✓[b] | ✓ | × | ✓ | Breast Cancer | ✓ (declined MV) | 6 | ND | Demyelinating | × | 1:3200/ 1:3200 | ✓ (red) | ✓ (yellow) | ✓ (red) | × | CYC | Death (Day 93) |
| P3 ($8^{th}$ / M) | CIDP | M&S | ✓[c] | × | × | × | Hodgkin lymphoma | ✓ | 5 | IgG-Lambda | Demyelinating | Axonal loss | 1:3200/ 1:1600 | ✓ (red) | ✓ (red) | ✓ (yellow) | ✓ (green) | ChIVPP | mRS 2 (1 year post RTX) |
| P4 ($7^{th}$ / M) | Atypical MMN | Motor only | ✓[d] | × | × | × | CLL | × | 5 | IgG-Lambda | Axonal | × | 1:800/ 1:1800 | ✓ (red) | ✓ (red) | ✓ (yellow) | ✓ (green) | CYC FDB | mRS 3 then 0 (6 then 9 months post RTX) |
| P5 ($8^{th}$ / M) | GBS | M&S | ✓[e] | ✓ | ✓ | × | No | ✓ | 5 | ND | Demyelinating | × | 1:400/ 1:800 | × | ✓ (yellow) | ✓ (yellow) | ✓ (green) | × | mRS 3 (6 months post RTX) |
| P6 ($5^{th}$ / M) | GBS | M&S | ✓[f] | ✓ | ✓ | ✓ | Nephrotic syndrome | ✓ | 5 | ND | Mixed | Axonal loss | 1:3200/ 1:6400 | ✓ (yellow) | ✓ (red) | ✓ (red) | ✓ (green) | × | mRS 1 (18 months post 1 cycle RTX, then clinical and serological relapse, successfully retreated) |
| P7 ($5^{th}$ / M) | GBS | M&S | ✓[b] | ✓ | × | × | No | ✓ | 6 | ND | × | × | 1:800/ 1:800 | × | ✓ (red) | × | × | × | Death (Day 18) |
| P8 ($8^{th}$ / M) | GBS | Motor only | ✓[g] | ✓ | ✓ | ✓ | Nephrotic syndrome | × (deemed unsuitable) | 6 | ND[h] | × | × | 1:400/ 1:400 | × | ✓ (red) | × | × | × | Death (Day 14) |

*Supplementary Table 2 - Clinical features of pan-neurofascin antibody positive patients*

**Key:**
✓ (red) Given but no response
✓ (yellow) Given but only partial and/ or transient response
✓ (green) Given with good response
× Not given / not present / not done

[a] II-VII, IX-XII including optic disc swelling
[b] Facial diplegia, subtle gaze palsy
[c] Bilateral III, IV and VI nerve palsies, facial diplegia
[d] Mild bulbar weakness
[e] Facial diplegia, dysphonia
[f] Facial diplegia, complete opthalmoplegia, optic disc swelling with reduced visual acuity, bulbar dysfunction
[g] Facial diplegia
[h] Clonal urinary lambda light chain

ND: Not detected
CN: Cranial Nerve
CLL: Chronic Lymphocytic Leukaemia
MV: Mechanical ventilation
mRS: modified Rankin Score
CYC: Cyclophsphamide
FDB: Fludarabine
IVIg: Intravenous Immunoglobulin
PLEx: Plasma Exchange
ChlVPP: Combination chemotherapy for Hodgkins Lymphoma (Chlorambucil, Vinblastine, Procarbazine, Predinisolone)

| Patient | Time from symptom onset | SNAPs | Motor Studies | | | | | | | | | | | | EMG | Neurophysiologist's report | Hadden classification | Rajabally classification |
|---|---|---|---|---|---|---|---|---|---|---|---|---|---|---|---|---|---|---|
| | | | Prolonged distal motor latency | | Slowed conduction velocity | | | Abnormal F-waves | | | Conduction block | | TD | CMAP | | | | |
| | | | >110 % | >150 % | <90% | <85% | <70% | >120 % | >150 % | absent | >0.7 | >0.5 | | | | | | |
| P1 | 2 weeks | Absent UL preserved sural | | | | | | | | | | | | Globally absent | No signs of acute denervation | Inexcitable | Inexcitable | Axonal GBS (inexcitable) |
| | 4 weeks | Globally absent | | | | | | | | | | | | Globally absent | Positive sharp waves and fibrillations | Inexcitable | Inexcitable | Axonal GBS (inexcitable) |
| | 8 weeks | Globally absent | | | | | | | | | | | | Globally absent | Florid denervation with no recordable motor units | Inexcitable | Inexcitable | Axonal GBS (inexcitable) |
| P2 | 4 weeks | Marginal reductions in amplitude and velocity | 1 | | 3 | 3 | 1 | 2 | | | 3 | 2 | | <50% LLN x 1 | ND | Demyelinating | Primary demyelinating | AIDP |
| P3 | 3 weeks [a] | Marginal reductions in amplitude and velocity | 2 | | 4 | 3 | | | | 8 | 7 | 4 | | <80% LLN x1 | Relative paucity of active denervation changes | Demyelinating | Primary demyelinating | AIDP |
| | 6 weeks [a] | Absent UL SNAPs | | | | | | | | | | | | Globally absent | Profuse acute denervation | Severe axonal loss | Inexcitable | Axonal GBS (inexcitable) |
| P4 | 4 weeks | Normal | 4 | | 2 | 2 | | 2 | | 6 | 4 | 4 | | <80% LLN x1 | | Conduction block | Primary demyelinating | AIDP |
| P5 | Day 5 | Mildly reduced UL amplitudes | 3 | 2 | 2 | 1 | | | | | 1 | | | <80% x6 <50% x 3 | No spontaneous or voluntary activity | Demyelinating | Primary demyelinating | AIDP |
| | 2 weeks | | 2 | | | | | | | | | | | <80% x1 <50% x 1 | Normal | Subtle demyelinating features, some improvement | Primary demyelinating | Axonal GBS |
| | 6 weeks | Absent / reduced | 4 | 4 | 1 | 1 | 1 | 1 | 1 | 1 | | | | <20% x4 <10% x3 | Pronounced active denervation | Secondary axonal loss | Primary demyelinating | AIDP |
| P6 | 3 weeks | Absent / reduced | 3 | 2 | 2 | 2 | 1 | | | 1 | 2 | 1 | | <50% x5 <10% x4 NR x3 | No spontaneous activity | Demyelinating | Primary demyelinating | AIDP |
| | 5 weeks | Globally absent | 1 | 1 | | | | | | | | | | <10% x4 NR x2 | Profuse positive sharp waves and fibrillations | Severe axonal loss | Primary axonal | Axonal GBS |

*Supplementary Table 3 - Detailed neurophysiological findings*

Numbers refer to how many individual nerves met the criteria stated. SNAP – sensory nerve action potential, CMAP – compound muscle action potential, AIDP – acute inflammatory demyelinating polyradiculoneuropathy, ND – not done. [a] Studies in this patient were performed 3 and 6 weeks after an acute neurological deterioration, which itself occurred after 4 months of insidiously progressive left leg weakness. Each study was assessed against the 1998 Hadden[4] and 2015 Rajabally[5] GBS electrodiagnostic criteria, as shown. Note the frequent presence of conduction block, absence of temporal dispersion, and frequent switch from demyelinating/AIDP to axonal classifications on repeat testing, suggestive of nodal pathology ("nodopathy") [6].

| Treatment | panNF (n=8) | | | | NF155 (n=15) | | | | Seronegative (n=194) | | | |
|---|---|---|---|---|---|---|---|---|---|---|---|---|
| | Number (%) treated | Response (n, % of treated) | | | Number (%) treated | Response (n, % of treated) | | | Number (%) treated | Response (n, % of treated) | | |
| | | Good | Partial or better | None or deterioration | | Good | Partial or better | None or deterioration | | Good | Partial or better | None or deterioration |
| **Steroids** | 4 (50%) | 0 (0%) | 1 (25%) | 3 (75%) | 14 (93%) | 3 (21%) | 8 (57%) | 6 (43%) | 62 | 5 (8%) | 26 (42%) | 36 (58%) |
| **IVIg** | 8 (100%) | 0 (0%) | 2 (25%) | 6 (75%) | 13 (87%) | 1 (8%) | 4 (31%) | 9 (69%) | 83 | 7 (8%) | 43 (52%) | 40 (48%) |
| **Plasma exchange** | 6 (75%) | 0 (0%) | 3 (50%) | 3 (50%) | 8 (53%) | 0 (0%) | 2 (25%) | 6 (75%) | 27 | 8 (30%) | 16 (59%) | 11 (41%) |
| **Rituximab** | 4 (50%) | 4 (100%) | 4 (100%) | 0 (0%) | 5 (33%) | 2 (40%) | 4 (80%) | 1 (20%) | 1 | 0 (0%) | 0 (0%) | 1 (100%) |

*Supplementary Table 4 - Physician reported treatment response by serological group*

IVIg – intravenous immunoglobulin. panNF – pan-neurofascin antibody positive, NF155 – neurofascin-155 antibody positive

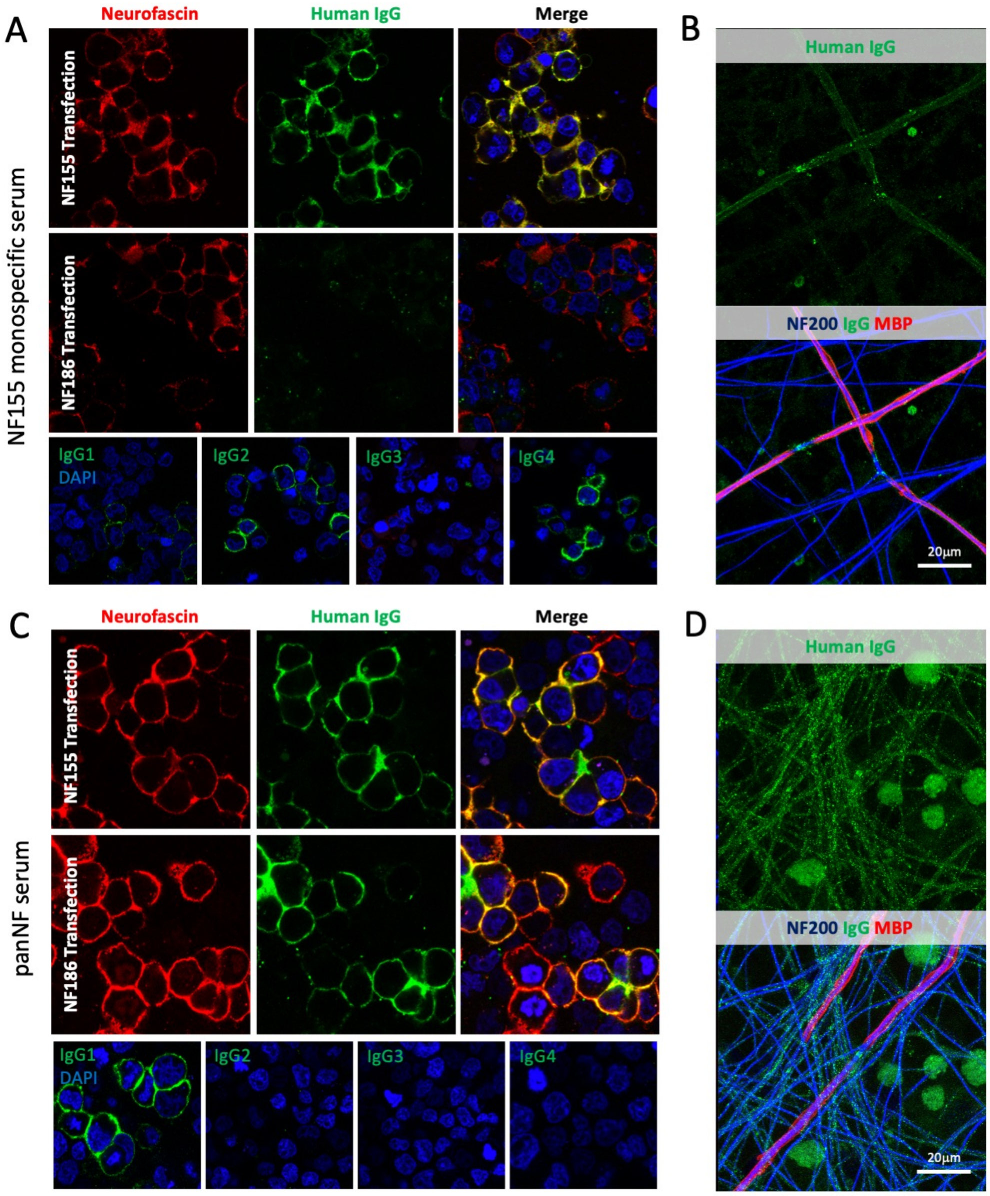

*Supplementary Figure 1 - Comparison between neurofascin-155 monospecific and pan-neurofascin serum reactivity on cell-based assays*

(A) A neurofascin(NF)-155 monospecific serum (upper panels) contains IgG which reacts exclusively with the membrane of NF155 and not NF186 transfected cells. IgG4, IgG2 and IgG1 subclass antibodies are present. (B) In myelinating co-cultures, IgG (green) is deposited over the outer surface of the myelin sheath (MBP, red) and at the node of Ranvier. (C) A panNF serum containing IgG reactive against both NF155 and NF186 transfected cells. Only IgG1 subclass antibodies with this reactivity are detected. (D) In myelinating co-cultures, IgG (green) is deposited along axons (NF200, neurofilament heavy, blue) and at the node of Ranvier.

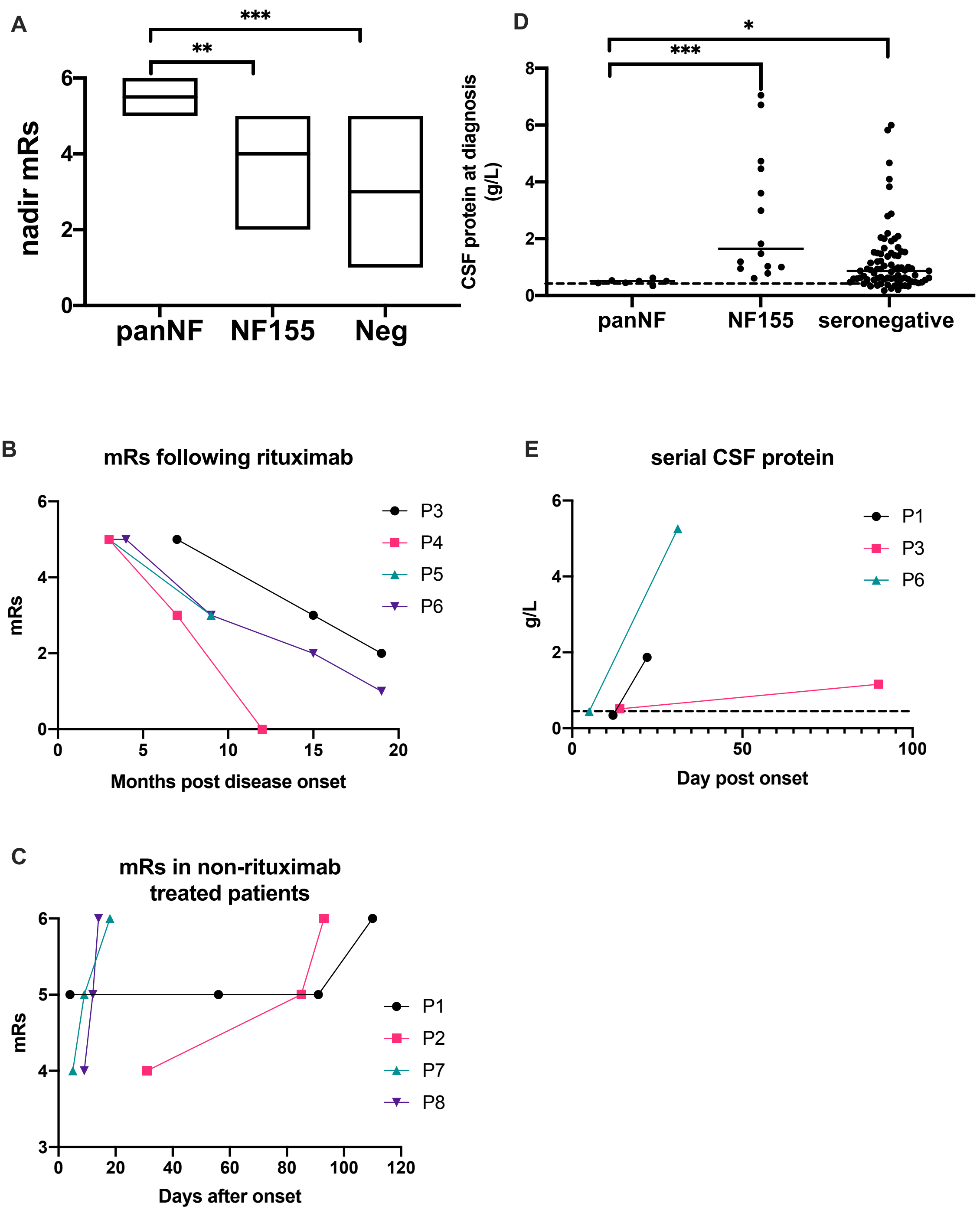

*Supplementary Figure 2 - Modified Rankin scores and CSF protein levels*

(A) The median nadir modified Rankin score (mRs) in the pan-neurofascin (panNF) antibody group was significantly higher than that in both the neurofascin-155 (NF155) antibody positive and seronegative groups. (B) Change in mRs in the 4 rituximab treated patients. The first datapoint for each patient represents the time of the first dose of rituximab. (C) Change in disability of non-rituximab treated patients. (D) CSF protein in the panNF group was either normal or only marginally elevated at diagnosis, and significantly lower than in both NF155 antibody positive and seronegative patients. (E) When performed, repeat CSF sampling showed abnormally increased protein levels at later time-points. Dashed lines indicate the upper limit of normal for CSF protein (0.45 g/L).

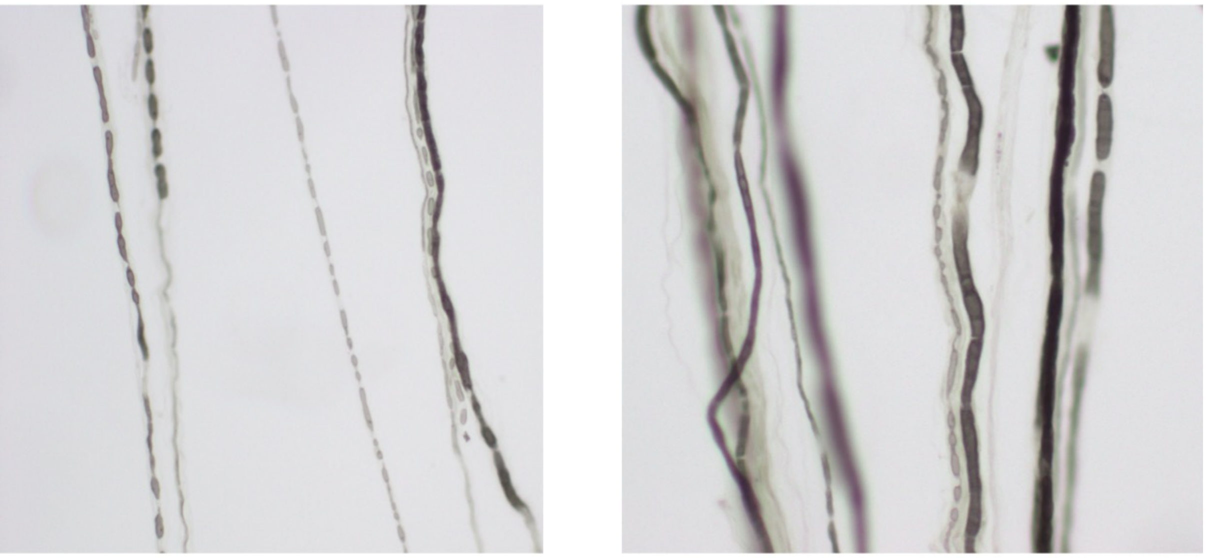

*Supplementary Figure 3 - Teased nerve fibres*

The Teased nerve fibres from the sural nerve biopsy of patient 6 show myelin ovoids consistent with axonal degeneration (left panel) and occasional short internodes (right panel).

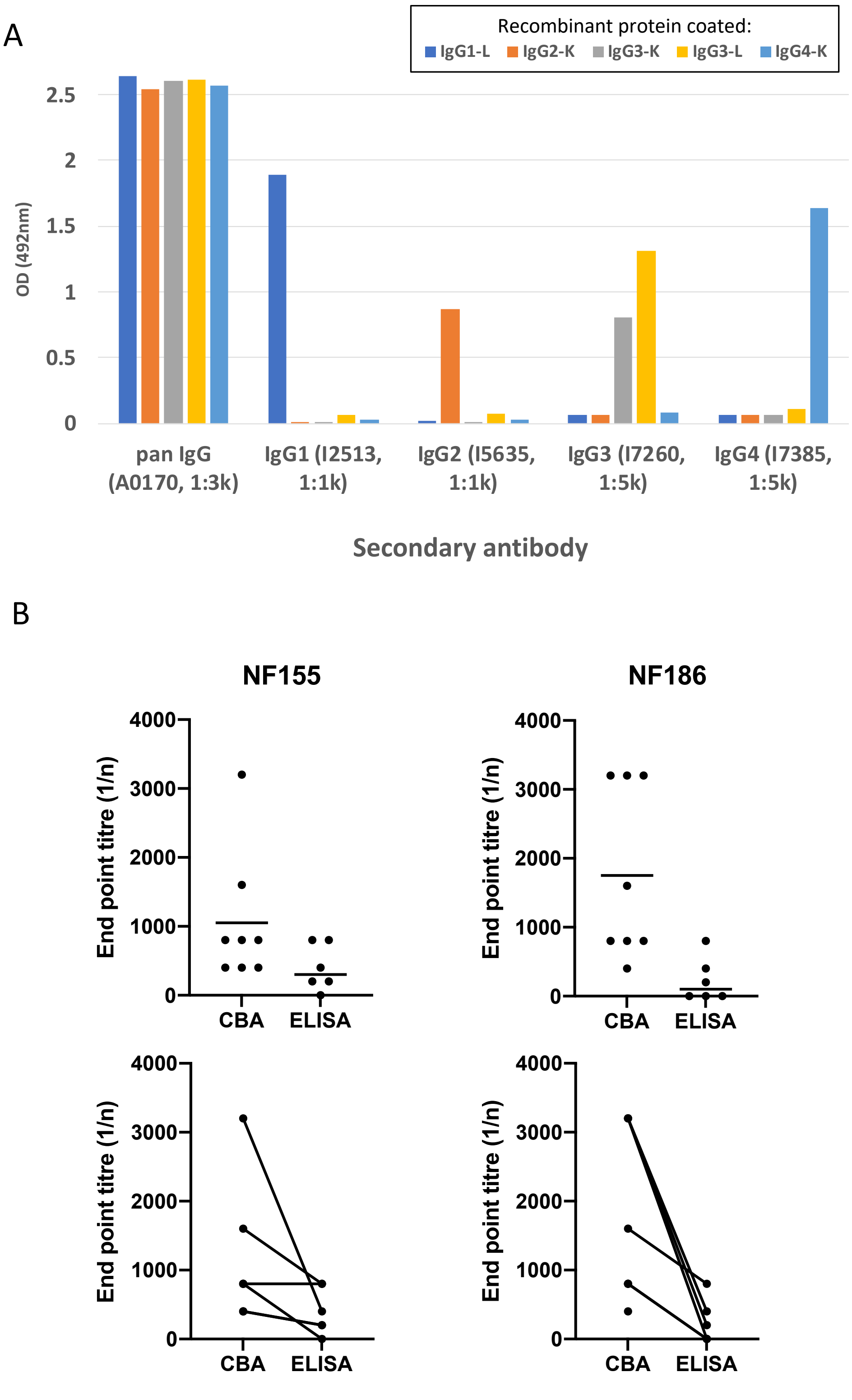

*Supplementary Figure 4 - Subclass specificity and comparison between CBA and ELISA*

(A) A pan-IgG secondary antibody detects all 5 human IgG recombinant proteins similarly. The subclass antibodies produce a signal only in wells coated with recombinants of their specified subclass. (B) End point titres on the cell-based assay (CBA) are consistently higher than by ELISA for both NF155 and NF186 antibodies. The upper panels show the individual data points and mean titre, the lower panels show the same data with lines linking individual patients' results across the 2 assays.

#### Online-only References

1 Delmont E, Manso C, Querol L, *et al.* Autoantibodies to nodal isoforms of neurofascin in chronic inflammatory demyelinating polyneuropathy. *Brain J Neurol* 2017;**140**:1851–8. doi:10.1093/brain/awx124

2 Fehmi J, Davies AJ, Antonelou M, *et al.* Contactin-1 Antibodies Link Autoimmune Neuropathies to Nephrotic Syndrome. Rochester, NY: : Social Science Research Network 2020. doi:10.2139/ssrn.3739819

3 Clark AJ, Kaller MS, Galino J, *et al.* Co-cultures with stem cell-derived human sensory neurons reveal regulators of peripheral myelination. *Brain J Neurol* 2017;**140**:898–913. doi:10.1093/brain/awx012

4 Hadden RDM, Cornblath DR, Hughes R a. C, *et al.* Electrophysiological classification of guillain-barré syndrome: Clinical associations and outcome. *Ann Neurol* 1998;**44**:780–8. doi:10.1002/ana.410440512

5 Rajabally YA, Durand M-C, Mitchell J, *et al.* Electrophysiological diagnosis of Guillain–Barré syndrome subtype: could a single study suffice? *J Neurol Neurosurg Psychiatry* 2015;**86**:115–9. doi:10.1136/jnnp-2014-307815

6 Uncini A, Susuki K, Yuki N. Nodo-paranodopathy: beyond the demyelinating and axonal classification in anti-ganglioside antibody-mediated neuropathies. *Clin Neurophysiol Off J Int Fed Clin Neurophysiol* 2013;**124**:1928–34. doi:10.1016/j.clinph.2013.03.025
